# The effect of pre-existing coronavirus antibodies on SARS-CoV-2 infection outcomes in exposed household members

**DOI:** 10.1101/2024.08.29.24312767

**Authors:** Ilse Westerhof, Reina Sikkema, Ganna Rozhnova, Janko van Beek, Marion Koopmans, Patricia Bruijning-Verhagen

## Abstract

**Background/Rationale:** We investigated the effect of pre-existing antibodies against SARS-CoV-2 and seasonal human coronaviruses on infection outcomes in Omicron BA1/2 exposed household members from January to March 2022.

**Methods:** Data from a prospective household study in the Netherlands were used including 63 households with 195 household members exposed to a SARS-CoV-2 Omicron BA1/2 index case. The protocol included repeated nose-throat swab and saliva RT-PCR testing, paired serology, and self-reported daily symptom scoring by household members. Infection outcomes included the occurrence of secondary infections, symptom severity, and CT-value trajectories. We studied the effect of baseline binding antibody levels for SARS-CoVs and seasonal coronaviruses (hCoV) NL63, 229E, HKU1 and OC43 spike protein, on SARS-CoV-2 infection outcomes.

**Results:** 132 of 195 (68%) exposed household members developed a SARS-CoV-2 infection. Among exposed household members, higher levels of SARS-CoV-2 and beta hCoV antibodies (HKU1 and OC43) at baseline, were associated with a reduced risk of secondary infection (adjusted Odds ratio 0.61; 95% Confidence interval 0.44-0.84 and aOR 0.72; 95%CI 0.56-0.92, respectively). No significant differences between antibody levels and symptom burden or CT-value trajectories were observed.

**Conclusions:** Our study suggests that prior SARS-CoV-2 and beta hCoV antibodies provide some protection against Omicron BA.1/BA.2 infection, while effects on symptom burden or CT-value could not be demonstrated. The results highlight the relatively limited, but not negligible role of cross-protective antibodies, especially when facing immune escape variants of SARS-CoV-2.

## Introduction

The emergence of the SARS-CoV-2 Omicron lineage was characterized by its notable immune escape from earlier variants and vaccines^1^. As new escape mutants continue to emerge, it is relevant to understand how pre-existing antibodies from earlier SARS-CoV-2 variants influence the risk of becoming infected, and if infection occurs, their influence on symptom burden, and CT-value trajectories. In addition, although the most immunodominant peptides of (seasonal) human coronaviruses (hCoVs) are quite distinct, antibodies to more conserved peptides may cross-react.

SARS-CoV-2 is classified as a beta coronavirus and most closely related to the seasonal beta coronaviruses hCoV-HKU1 and hCoV-OC43, and therefore more distantly related to seasonal alpha coronaviruses hCoV-229E and hCoV-NL63. Research indicates infections with hCoVs occur repeatedly throughout life, with varying infection rates and immunity waning across age groups^2,3^. For SARS-CoV-2, a stable epidemiological pattern has not been established yet, but endemic patterns with recurring infections have been observed^4,5^.

The role of cross-reactive SARS-CoV-2 and hCoV antibodies in the complex interplay between reinfection patterns, immunity and virus evolution remains to be determined^6,7^. While there is a growing body of research indicating that SARS-CoV-2 antibodies can be cross-protective between different variants^3^, evidence on cross-protection from antibodies against other hCoVs is inconsistent^8–10^.

Here, we leverage data from a detailed household transmission study conducted during the first Omicron wave in early 2022 to explore the effect of pre-existing antibodies to SARS-CoV-2 and to the four seasonal hCoVs on Omicron infection risk in exposed household members, and on infection outcomes including symptom severity and CT-value trajectories.

## Methods

### Study design and study population

The VERDI-RECOVER household study was conducted in the Netherlands from January to March 2022 during the emergence of the Omicron BA.1/BA.2. The study protocol has been described elsewhere ^11^. In brief, households were eligible following a positive SARS-CoV-2 test result, either by rapid antigen detection assay or RT-PCR, in the household index case, and no positive test result among any of the other household members in the previous two weeks. Symptoms of household members were monitored on a daily basis by means of a symptom diary until 21 days after the last symptom onset. Study participants were asked to self-collect nose-throat swab (NTS) and saliva samples on days 0, 2, 4, 7, and 14, and upon onset of new COVID-19-related symptoms. Paired dried blood spots (DBS) were collected at enrollment and at least 4 weeks after enrollment or 10 days after the last symptom-onset. Additional data collection included COVID-19 vaccination status, SARS-CoV-2 infection history, and medical comorbidities, and was done using a questionnaire.

#### Diagnostic testing

NTS and saliva samples were tested for SARS-CoV-2 by RT-PCR, using the TaqPath™ COVID-19 ORF1, N and S gene targeted RT-PCR kit on a QUANTSTUDIO™5 (Thermofisher Scientific).

To investigate the presence of antibodies, DBS were tested in a final dilution of 1:40 for immunoglobin G antibodies reactive with the target antigens using a multiplex protein microarray. Antibody binding was measured for the following human coronavirus targets: NL63 nucleocapsid protein (NP), S1 subunit, and S ectodomain; 229E NP and S1; HKU NP and S1; OC43 S ectodomain; as well as SARS-CoV-2 S1, NP, and S ectodomain. Recombinant spike proteins of S1 or the S ectodomain were expressed in HEK293 cells NP proteins were produced in Escherichia coli (for hCoV-OC43, hCoV-229E, hCoV-NL63, and hCoV-HKU1; Medix Biochemica, Espoo, Finland) or insect cells and baculovirus (SARS; Sino Biological, Beijing, China)^12^. More details about the laboratory process are given elsewhere^13^.

The study was reviewed and ethically approved by the Medical Ethical committee Utrecht, The Netherlands (reference number 17-069/M). Written informed consent was obtained from all participating household members and/or their legal representatives.

For these analyses we selected all household members, excluding the index case, for which results from PCR testing of NTS and the baseline DBS sample results were available.

### Definitions

SARS-CoV-2 infection in household members was defined as a positive RT-PCR result, with a cycle threshold (Ct) value 40 and lower, in at least one of the serial NTS or saliva samples. A household member was identified as a coprimary case when the NTS collected on the day of enrollment tested positive for SARS-CoV-2 RNA. Subsequently, a consecutive period with CT-values below 40 was defined as the duration of virus positivity. From this period, we also marked the duration of CT-values below 30 as a proxy for the infectious period. The peak in viral load was determined as the lowest detected CT-value during the infection.

The presence of a previous SARS-CoV-2 infection prior to study enrolment was based on either a reported prior positive SARS-CoV-2 test result or on the presence of SARS NP antibodies at baseline.

Antibody status for SARS-CoV-2 and seasonal hCoV was assessed based on DBS samples at baseline. As antibody measurements based on fluorescence signals derived from the microarray on DBS samples were right censored and no protective reference levels are known, we used a non-parametric approach for analysis: for each antibody target, we grouped antibodies into ‘high’ or ‘low’ relative to the median value of the study population. Next, we calculated a cumulative score of high antibodies per coronavirus (SARS-CoV-2, hCoV-NL63, hCoV-229E, hCoV-HKU, hCoV-OC43) and for seasonal coronaviruses grouped as alpha or beta. This cumulative antibody score per coronavirus included could vary between 0 and 3 depending on the number of recombinant viral antigens (spike ectodomain, spike S1, nucleoprotein) for which the antibody titer was above the median of the study population. For hCoV-OC43, only one antibody target was included, and no cumulative score was calculated. Similarly, the cumulative antibody scores for alpha coronaviruses varied between 0 to 4, and for beta coronaviruses between 0 and 6, depending on the number of recombinant viral antigens (spike ectodomain, spike S1, and nucleoprotein) for which the antibody titer was above the median of the study population.

Cross-reactive antibodies are antibodies that are generated in response to one virus, such as SARS-CoV, and can also bind to antigens from another virus, such as SARS-CoV-2. Cross-protection occurs if these antibodies reduce success of subsequent infection. We use these definitions to assess how immune responses of different coronaviruses interact and contribute to the analysis of antibody levels and Omicron infection risk, symptom severity and CT-value trajectories.

SARS-CoV-2 disease severity was categorized into symptomatic disease, pauci-symptomatic, and asymptomatic episodes. Symptomatic disease was defined as: 1) onset of fever OR 2) two consecutive days with one respiratory (cough, sore throat, runny or congested nose, dyspnea) and one systemic symptom (headache, muscle ache, sweats or chills or tiredness) or with at least two respiratory symptoms. An episode was defined as pauci-symptomatic if symptoms occurred within the specified time-window but remained below the threshold for a symptomatic disease episode, and asymptomatic if no symptoms were reported. Subjects meeting the criteria for symptomatic disease additionally received a daily symptom severity score which consisted of a 5-point Likert scale per reported symptom present, except for fever, which was categorized as <38/38-39/39-40/>40 degrees Celsius. The cumulative severity score was defined as the sum of daily scores reported during the symptomatic period. We used the daily symptom data and date of positive test results to define the onset and resolution of a SARS-CoV-2 episode and to calculate the episode duration. An episode started on the day of symptom onset, which had to fall within the seven days before to seven days after the first positive test result. An episode ended on the last symptomatic day that was followed by at least two days without any symptoms.

### Statistical analyses

We employed a combination of parametric and non-parametric descriptive statistics to summarize household member characteristics and pre-existing antibody profiles. We assessed correlations between cumulative antibody scores for different coronaviruses within an individual at baseline using Pearson correlation coefficients. This analysis aimed to identify potential correlations between antibodies for SARS-CoV-2 and seasonal hCoVs.

The impact of the cumulative antibody score for alpha and beta coronaviruses and for each individual coronavirus on the SARS-CoV-2 infection risk was evaluated in multivariable analyses with Generalized Linear Models (GLMs). Results are presented as adjusted Odds Ratios (aORs) and 95% confidence intervals (95% CI) per one level increase in cumulative antibody score, and reflect the relative effect of high versus low antibodies at baseline.

For the subgroup of SARS-CoV-2 infected household members, we summarized disease outcomes including disease category, cumulative severity score, episode duration, as well as CT-value trajectories including duration of PCR positivity, duration of CT-value < 30, and lowest CT-value detected. The impact of baseline antibodies to the different coronaviruses on disease outcomes and on CT-value trajectories was evaluated using GLMs. Results are presented as mean difference with 95% CI and represent the effect of low versus high antibodies on disease and CT-value outcomes.

All models were adjusted for age of household member and age of the index case. For analyses excluding betaviruses and SARS-CoV-2, an additional adjustment was made for cumulative SARS-CoV-2 antibody score at baseline to account for the potential influence of cross-reactive antibodies. First, we assessed the effect of cumulative antibody scores for SARS-CoV-2 and for each seasonal hCoV. Next, any potential association, defined as a p-value <0.1, was further explored in a stratified analysis of each antibody target separately, that was included in the cumulative score.

We performed a sensitivity analysis by excluding co-primary cases from the analyses, as the date of infection cannot be established for these subjects and therefore the baseline sample may not exclusively reflect pre-existing antibodies.

All statistical analyses were performed using R version 4.0.3 (R Core Team, 2020). The significance level was set at alpha=0.05.

### Code Availability

The codes reproducing the results of this study are publicly available at https://github.com/IWesterhof/Antibody-profiles-and-COVID-19.

### Data Availability

The data analyzed for this manuscript are not publicly available due to privacy concerns. The data contain sensitive personal information, and sharing these data could compromise participant confidentiality. For more information, please contact the corresponding author.

## Results

In total, 195 household members were included in the analyses with a median age of 39 years. In total, 48.7% (n=95) were female, and 70.8% (n=138) had received at least the primary series of COVID-19 vaccination. The majority of the household index cases were aged <12 years (58.3%). Baseline characteristics and antibodies for all household members are shown in Table 1. Weak to moderate positive correlations were identified between cumulative antibody scores for SARS-CoV-2 and seasonal hCoV at baseline and end of follow-up, with correlation coefficients ranging from 0.2 to 0.37 (p<0.01; Supplementary Figure 1), suggesting limited cross-reactivity between viral antigens.

**Table 1.** Characteristics of SARS-CoV-2 exposed household members.

|  |  | Median (IQR) / n (%) |  |  |  |
| --- | --- | --- | --- | --- | --- |
|  |  | All (n=195) | SARS-CoV-2 Infected (n=132) | Not infected (n=63) | P-value |
| Age |  | 39 (11, 45) | 36 (10, 43) | 43 (16.5, 47) | <0.001 |
| Vaccination status |  |  |  |  | 0.008 |
| Not vaccinated |  | 51 (26.2%) | 43 (32.6%) | 8 (12.7%) |  |
| Incomplete Primary |  | 6 (3.1%) | 2 (1.5%) | 4 (6.3%) |  |
| Primary series |  | 55 (28.2%) | 35 (26.5%) | 20 (31.7%) |  |
| Booster |  | 83 (42.6%) | 52 (39.4%) | 31 (49.2%) |  |
| Prior SARS-CoV-2 infection |  | 95 (48.7%) | 58 (43.9%) | 37 (58.7%) | 0.053 |
| Age household index case |  |  |  |  | 0.017 |
| <12 |  | 112 (58.3%) | 84 (64.6%) | 28 (45.2%) |  |
| 12-17 |  | 53 (27.6%) | 28 (21.5%) | 25 (40.3%) |  |
| ≥18 |  | 27 (14.1%) | 18 (13.8%) | 9 (14.5%) |  |
| Unknown |  | 3 | 2 | 1 |  |
| Baseline hCoV antibody levels |  | Median (IQR) | Median (IQR) | Median (IQR) | P-value |
| SARS-CoV-2 | Ecto | 60,942 (59,145, 61,977) | 60,772 (3,867, 61,640) | 61,673 (60,472, 62,152) | <0.001 |
|  | NP | 963 (664, 1,742) | 937 (675, 1,421) | 1,249 (595, 3,965) | 0.2 |
|  | S1 | 60,845 (57,349, 61,824) | 60,218 (3,526, 61,690) | 61,320 (60,429, 62,101) | <0.001 |
| NL63 | Ecto | 61,177 (60,114, 62,074) | 61,206 (60,124, 62,166) | 61,150 (59,978, 61,983) | 0.7 |
|  | NP | 28,477 (14,104, 60,481) | 26,609 (13,290, 59,651) | 48,947 (16,095, 61,154) | 0.031 |
|  | S1 | 6,038 (2,112, 9,790) | 5,698 (1,879, 9,784) | 6,346 (3,144, 9,921) | 0.4 |
| 229E | NP | 24,698 (11,363, 60,420) | 22,301 (9,966, 60,272) | 29,668 (15,942, 60,420) | 0.073 |
|  | S1 | 61,717 (60,108, 62,402) | 61,609 (59,167, 62,372) | 61,899 (61,370, 62,484) | 0.038 |
| HKU1 | NP | 6,670 (4,772, 10,201) | 6,585 (5,024, 9,823) | 7,108 (4,704, 11,080) | 0.7 |
|  | S1 | 57,723 (21,308, 60,964) | 54,910 (19,654, 60,666) | 59,763 (27,269, 61,343) | 0.026 |
| OC43 | Ecto | 57,264 (54,158, 59,901) | 57,706 (54,455, 59,970) | 56,904 (53,826, 59,605) | 0.6 |
| Antibody above 67,000 remaining unassessed due to right-censoring. |  |  |  |  |  |

### Infection risk

In total, 132 household members (67.7%) were infected with SARS-CoV-2 during the follow-up (Table 1). Of these, 65 household members (49.2%) were considered to be a coprimary case. Infected household members were younger than non-infected household members (age 36 versus age 43 years, P<0.001), and more frequently unvaccinated (32.6% versus 12.7%, P=0.005). The absolute pre-existing antibody titers for SARS-CoV-2 spike ectodomain and spike S1 were significantly higher in uninfected compared to infected household members (P<0.001 and P<0.05, respectively). Individuals with a reported prior infection had significantly more frequent ‘high’ NP antibodies (85.0% among infected versus 16.0% among uninfected, P<0.001), but not significantly higher S1 and S ectodomain antibodies (both P=0.5). Additionally, hCoV-NL63 NP, hCoV-229E spike S1, and hCoV-HKU1 spike S1 antibody titers were also found to be significantly higher in uninfected household members (all P<0.05; Supplementary Figure 2). In multivariable analysis, higher cumulative beta coronavirus and SARS-CoV-2 antibody scores at baseline were significantly associated with a reduced risk of infection (aOR 0.72; 95%CI 0.56-0.92 and 0.61; 95%CI 0.44-0.84). Stratified analyses per viral antigen showed a significant protective effect of SARS-CoV-2 spike ectodomain (aOR 0.44; 95%CI 0.22-0.84) and of SARS-CoV-2 spike S1 (aOR 0.42; 95%CI 0.21-0.81) antibodies. In adjusted analysis, the effect of cumulative antibodies against separate seasonal hCoVs on the risk of SARS-CoV-2 infection was no longer significant (Table 2).

**Table 2.**
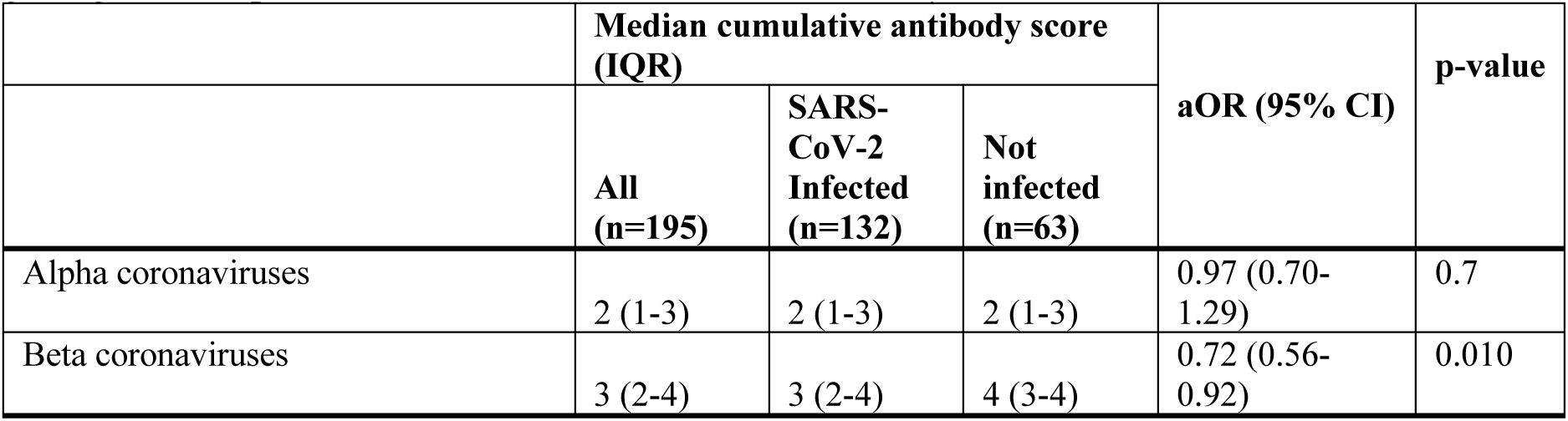
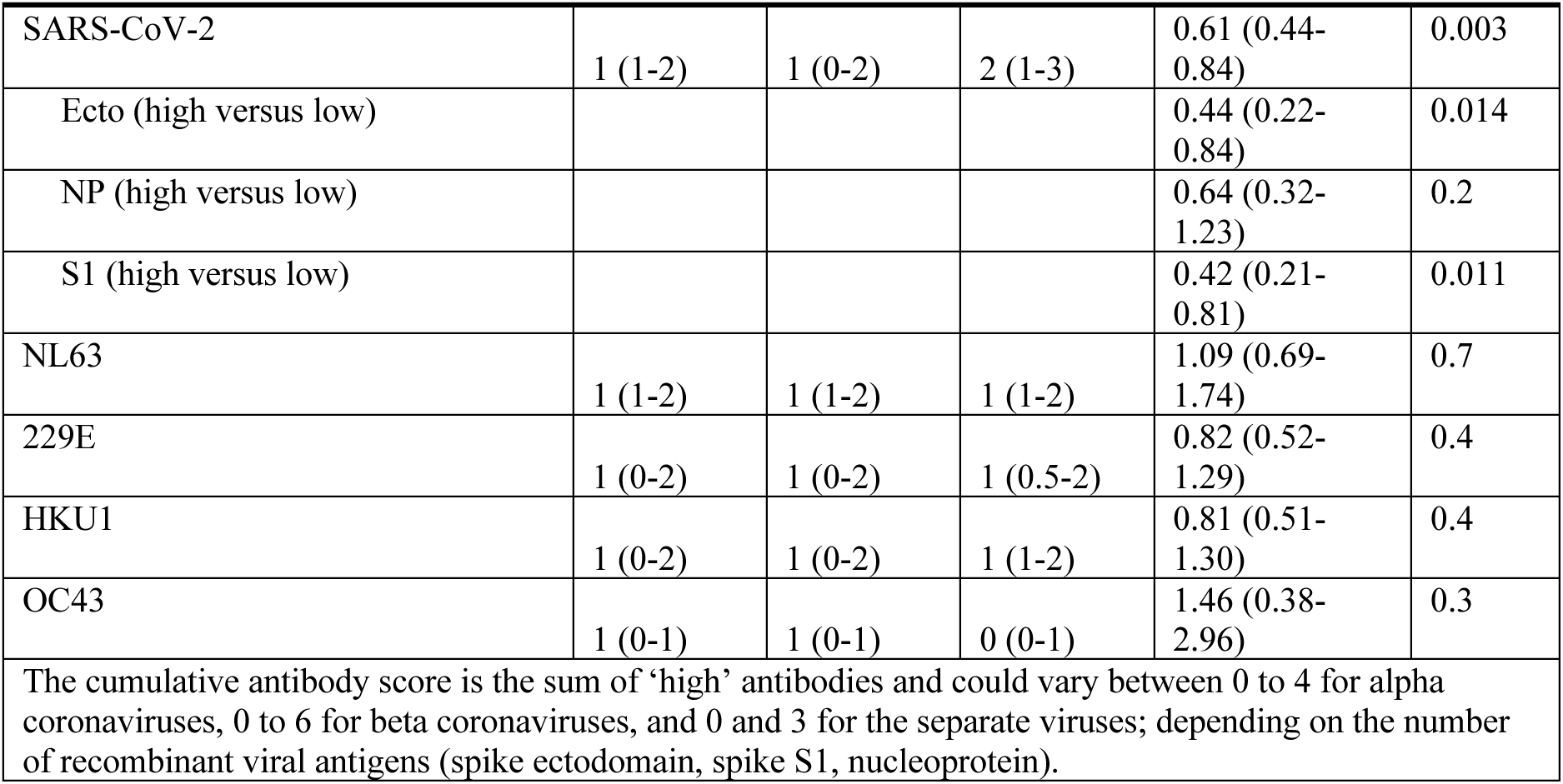
SARS-CoV-2 infection risk by baseline antibody status for different coronaviruses; aORs for getting infected per one level increase in cumulative antibody score.

|  | Median cumulative antibody score (IQR) |  |  | aOR (95% CI) | p-value |
| --- | --- | --- | --- | --- | --- |
|  | All (n=195) | SARS-CoV-2 Infected (n=132) | Not infected (n=63) |  |  |
| Alpha coronaviruses | 2 (1-3) | 2 (1-3) | 2 (1-3) | 0.97 (0.70-1.29) | 0.7 |
| Beta coronaviruses | 3 (2-4) | 3 (2-4) | 4 (3-4) | 0.72 (0.56-0.92) | 0.010 |
| SARS-CoV-2 | 1 (1-2) | 1 (0-2) | 2 (1-3) | 0.61 (0.44-0.84) | 0.003 |
| Ecto (high versus low) |  |  |  | 0.44 (0.22-0.84) | 0.014 |
| NP (high versus low) |  |  |  | 0.64 (0.32-1.23) | 0.2 |
| S1 (high versus low) |  |  |  | 0.42 (0.21-0.81) | 0.011 |
| NL63 | 1 (1-2) | 1 (1-2) | 1 (1-2) | 1.09 (0.69-1.74) | 0.7 |
| 229E | 1 (0-2) | 1 (0-2) | 1 (0.5-2) | 0.82 (0.52-1.29) | 0.4 |
| HKU1 | 1 (0-2) | 1 (0-2) | 1 (1-2) | 0.81 (0.51-1.30) | 0.4 |
| OC43 | 1 (0-1) | 1 (0-1) | 0 (0-1) | 1.46 (0.38-2.96) | 0.3 |
| The cumulative antibody score is the sum of 'high' antibodies and could vary between 0 to 4 for alpha coronaviruses, 0 to 6 for beta coronaviruses, and 0 and 3 for the separate viruses; depending on the number of recombinant viral antigens (spike ectodomain, spike S1, nucleoprotein). |  |  |  |  |  |

### Disease outcomes

Of the 132 infected household members, 63.6% (n=84) were symptomatic, 25.8% (n=34) pauci-symptomatic, and the remaining 14 subjects were asymptomatic (10.6%). Among those symptomatic, the mean cumulative severity score was 76.4 (SD 71.8), and the mean symptom duration among symptomatic and pauci-symptomatic subjects, was 13.8 (SD 11.1) and 5.9 days (SD 5.6), respectively. High levels of NP antibodies for SARS-CoV-2, hCoV-NL63, and hCoV-229E were more frequent in asymptomatic and pauci-symptomatic subjects (Suppl Figure 3). In multivariable analysis, no significant associations were found between any of the SARS-CoV-2 or hCoV cumulative antibody scores and disease severity. Similarly, the cumulative antibody scores for any of the coronaviruses were not significantly associated with the cumulative severity score or illness duration, except for higher cumulative hCoV-NL63 antibodies, which were associated with an 8.4 increase in cumulative severity score per one level increase in cumulative antibody score (P=0.030; Table 3). Additional analyses of the different hCoV-NL63 antibody targets did not show significant associations with disease outcomes.

**Table 3.** Symptomatology of SARS-CoV-2 infection by baseline antibody status for different coronaviruses; aORs and adjusted mean difference per one level increase in cumulative antibody score.

|  | Disease severity |  |  |  | Cumulative severity score |  | Symptom duration in days |  |
| --- | --- | --- | --- | --- | --- | --- | --- | --- |
|  | Symptomatic | Pauci-symptomatic | Asymptomatic | p-value | mean difference (95% CI) | p-value | mean difference (95% CI) | p-value |
| n (%) / Mean (SD) | 84 (63.6%) | 34 (25.8%) | 14 (10.6%) |  | 76 (72) |  | 11 (11) |  |
| Alpha coronaviruses | Reference | 1.22 (0.84-1.79) | 1.01 (0.59-1.74) | 0.6 | 13 (0.29-27) | 0.055 | -0.32 (-2.1-1.5) | 0.7 |
| Beta coronaviruses | Reference | 0.96 (0.69-1.32) | 1.23 (0.05-0.71) | 0.6 | 8.3 (4.3-21) | 0.2 | 0.42 (-1.2-2.0) | .6 |
| SARS-CoV-2 | Reference | 0.97 (0.62-1.52) | 1.40 (0.77-2.56) | 0.51 | 5.7 (-11-22) | 0.5 | -0.47 (-2.6-1.6) | 0.7 |
| NL63 | Reference | 1.20 (0.67-2.15) | 0.71 (0.32-1.54) | 0.5 | 8.4 (0.84-16) | 0.030 | -0.45 (-1.5-0.59) | 0.4 |
| 229E | Reference | 1.31 (0.75-2.31) | 1.47 (0.67-3.22) | 0.5 | 11 (-9.9-32) | 0.3 | -0.22 (-2.9-2.4) | 0.9 |
| HKU1 | Reference | 0.81 (0.45-1.48) | 1.78 (0.79-4.01) | 0.23 | 11 (-11-33) | 0.3 | 1.3 (-1.5-4.0) | 0.4 |
| OC43 | Reference | 1.36 (0.53-3.45) | 0.26 (0.07-1.0) | 0.07 | 7.4 (-25-39) | 0.6 | 1.7 (-2.4-5.8) | 0.4 |
| The cumulative antibody score is the sum of 'high' antibodies and could vary between 0 to 4 for alpha coronaviruses, 0 to 6 for beta coronaviruses, and 0 and 3 for the separate viruses; depending on the number of recombinant viral antigens (spike ectodomain, spike S1, nucleoprotein). |  |  |  |  |  |  |  |  |

### CT-value trajectories

Among infected household members, the mean duration of RT-PCR positivity in NTS samples was 9 days (SD 5.5), while CT-values <30 were present for a mean of 7 days (SD 4.3, table 4). The mean peak in viral load corresponded to a CT-value of 18.9 (SD 5.6) two days after onset symptoms. NTS samples had significantly lower CT-values compared to saliva samples (Figure 1A). Lower CT-values correlated with higher disease severity categories (Figure 1B; P<0.001). There were no differences in duration of positivity (CT<40), duration of CT<30, and peak viral load between subjects with high versus low cumulative antibody scores for SARS-CoV-2 or any of the hCoVs, which was confirmed in multivariable analysis (Figure 2 and Table 4).

**Fig. 1.**
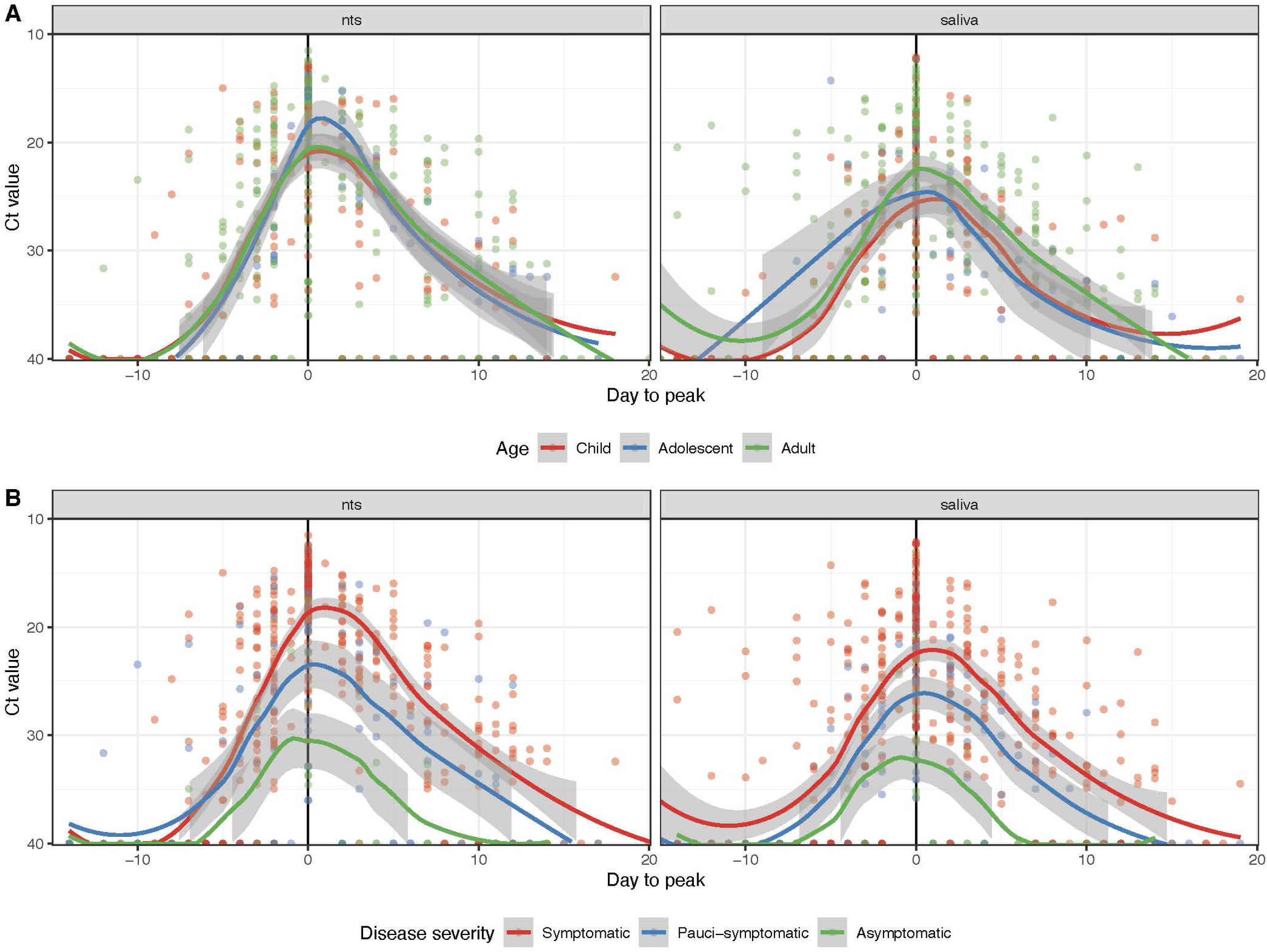
CT-value trajectories for SARS-CoV-2 infected individuals. The observed viral load peak is set at day 0 for each subjects A) CT-value trajectories by age group (red:Child < 12 yo; Blue: Adolescent 12-17 yo; Green: Adult > 18 yo) and sample type, B) CT-value trajectories by disease severity (Red: Symptomatic; Blue: Pauci-symptomatic; Green: Asymptomatic) and sample type. Lines represent LOESS smoothed values with 95% confidence intervals.

**Fig. 2.**
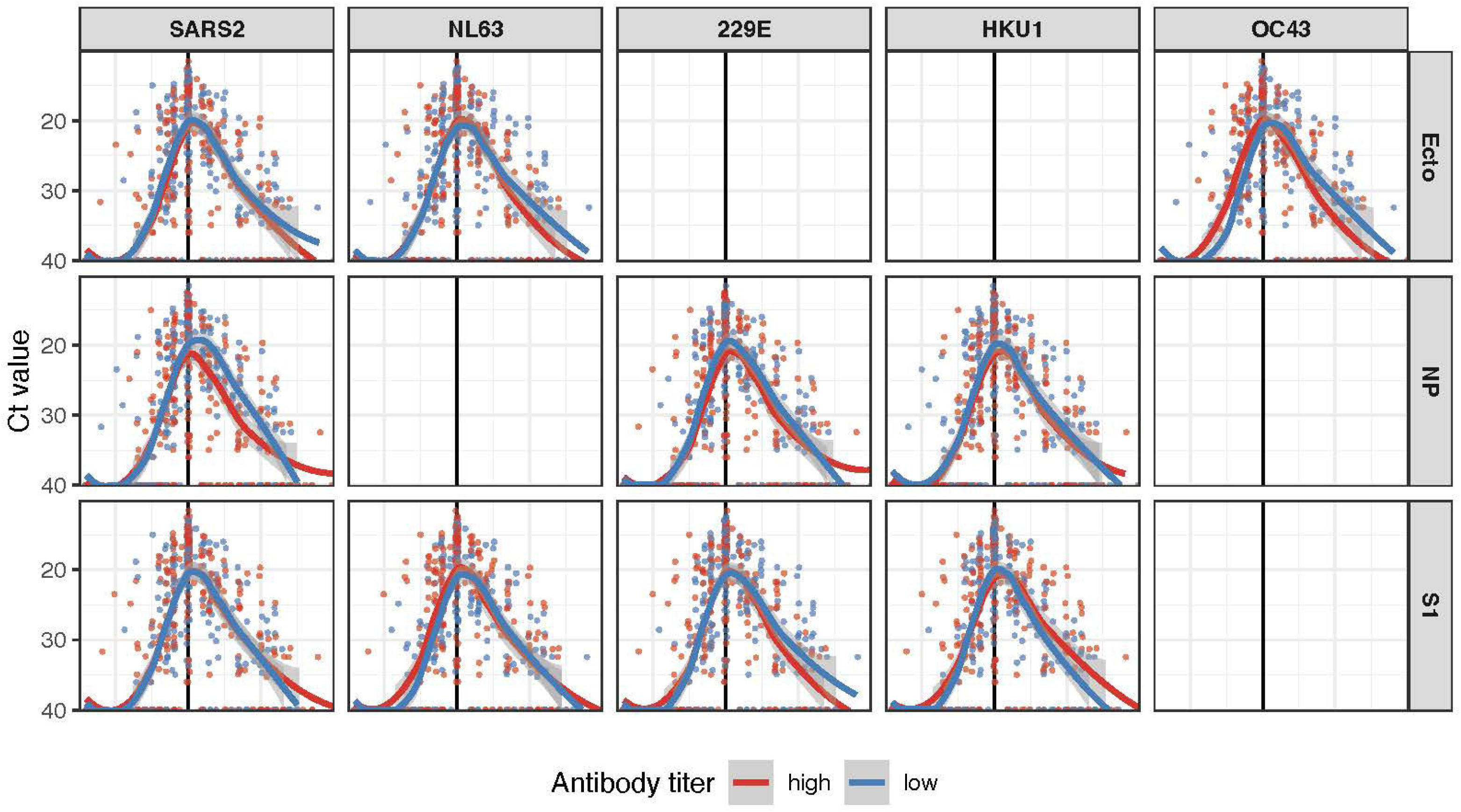
CT-value trajectories for SARS-CoV-2 infected individuals by baseline antibody status for different coronaviruses (SARS-CoV-2, NL63, 229E, HKU1, OC43). Antibody titers are defined as high (above the population median; red lines) and low (below the population median; blue lines) and shown per antibody target. Lines represent LOESS smoothed values with 95% confidence intervals.

**Table 4.** CT-value trajectories in SARS-CoV-2 infected subjects by baseline antibody status for different coronaviruses; adjusted mean difference per one level increase in cumulative antibody score.

|  | Duration of positivity (CT<40) |  | Duration of CT<30 |  | Peak viral load (Lowest CT-value) |  |
| --- | --- | --- | --- | --- | --- | --- |
|  | Mean difference (95% CI) | p-value | Mean difference (95% CI) | p-value | Mean difference (95% CI) | p-value |
| n (%) / Mean (SD) | 9.4 (5.5) |  | 7 (4.3) |  | 18.9 (5.6) |  |
| Alpha coronaviruses | -0.25 (-.12-0.68) | 0.6 | -0.18 (-0.91-0.56) | 0.6 | -0.44 (-1.4-0.49) | 0.4 |
| Beta coronaviruses | -0.20 (-0.99-0.58) | 0.6 | -0.41 (-1.1-2.3) | 0.2 | 0.24 (-0.55-1.0) | 0.6 |
| SARS-CoV-2 | -0.19 (-1.2-0.86) | 0.7 | -0.48 (-1.3-0.35) | 0.3 | 0.31 (-0.75-1.4) | 0.6 |
| NL63 | 0.5 (-1.3-1.5) | 0.9 | 0.21 (-0.88-1.3) | 0.7 | -1.1 (-2.5-0.26) | 0.12 |
| 229E | -0.67 (-2.1-0.71) | 0.3 | -0.61 (-1.7-0.48) | 0.3 | 0.14 (-1.3-1.5) | 0.8 |
| HKU1 | -0.03 (-1.5-1.4) | >0.9 | -0.11 (-1.3-1.1) | 0.9 | 0.75 (-0.69-2.2) | 0.3 |
| OC43 | -0.56 (-2.7-1.5) | 0.6 | -0.49 (-2.2-1.2) | 0.6 | -1.3 (-3.4-0.84) | 0.2 |
| The cumulative antibody score is the sum of 'high' antibodies and could vary between 0 to 4 for alpha coronaviruses, 0 to 6 for beta coronaviruses, and 0 and 3 for the separate viruses; depending on the number of recombinant viral antigens (spike ectodomain, spike S1, nucleoprotein). |  |  |  |  |  |  |

### Sensitivity analyses infection risk

After excluding 65 (49.2%) coprimary cases, the effect of cumulative antibodies on infection risk did not change (data not shown).

## Discussion

In this household transmission study conducted during the first Omicron wave in early 2022, we observed an overall SARS-CoV-2 infection rate of 67.7% among the 195 exposed household members. Our study confirmed that higher baseline levels of SARS-CoV-2 antibodies were associated with lower odds of infection (aOR 0.61; 95%CI 0.44-0.84). Moreover, cumulative high levels of antibodies against seasonal beta hCoVs significantly alters the risk of infection (aOR 0.72; 95%CI 0.56-0.92), but this effect was no longer significant when stratified by separate seasonal coronaviruses. Our study further indicates a potential effect of antibodies specific against NP of SARS-CoV-2, hCoV-NL63, and hCoV-229E. Higher NP antibodies were present in asymptomatic and pauci-symptomatic subjects, compared to symptomatic subjects, while for target S1 and ecto, such antibody differences were not observed. The lack of an effect from S1 and ecto antibodies may have diluted the association in multivariate analyses between the cumulative antibody scores, which combines the different targets into one score, and symptom burden. NP antibodies are markers of infection rather than vaccination, supporting the idea that hybrid immunity provides enhanced defense against severe symptoms.^14^ No effect of baseline coronavirus antibodies on SARS-CoV-2CT-value trajectories was observed. Our study found weak to moderate positive correlations between the number of high SARS-CoV-2 and seasonal hCoV antibodies. This suggests limited cross-reactivity between viral antigens.

The results indicate that higher SARS-CoV-2 binding antibody levels, whether from vaccination or past infection, are cross-protective against infection with a different SARS-CoV-2 variant. This is in line with observations earlier during the pandemic when presence of antibodies against SARS-CoV-2 pre-Delta variant was found to be protective against infection with Delta variant^15,16^. Also, our study did find a significant protective effect of high levels of seasonal beta hCoV antibodies (SARS-CoV-2, HKU1, and OC43) against SARS-CoV-2 infection, but not for the separate seasonal hCoV’s. This is in line with the observations from Sermet-Gaudelus et al^8^, who found no significant differences in baseline hCoV antibody titers among SARS-CoV-2 infected versus uninfected children during the first year of the pandemic. However, some other studies report a non-significant trend towards cross-protection, including a longitudinal study conducted in the first year of the pandemic for baseline hCoV N antibodies^17^.

Our study supports an attenuating effect of prior infection with SARS-CoV-2, but also hCoV NL63 and 229E on symptom burden of SARS-CoV-2 BA.1/BA.2 infection, as suggested by the higher antibodies against SARS-CoV-2 NP, hCoV-NL63 NP, and hCoV-229E NP in asymptomatic and pauci-symptomatic, compared to symptomatic infections in the univariate analyses. A similar effect from S1 antibodies, as found in a cross-sectional exploratory study conducted in 2020 with 165 subjects, associating hCoV-229E spike S1 antibodies with the development of milder COVID-19 symptoms or asymptomatic cases, was not observed^9^. Some studies also suggest a direct correlation between pre-existing antibodies against hCoV-OC43 and COVID-19 disease severity. An early pandemic study in China found a correlation between COVID-19 severity and hCoV-OC43 spike protein antibodies^18^, while a study in France and Germany showed lower hCoV-OC43 NP antibodies for critically ill patients^19^. The difference in findings may lie in variations in patient populations, clinical settings, and viral variants, as our study only included non-hospitalized patients.

In this study, we could not demonstrate an effect of pre-existing SARS-CoV-2 or hCoV antibodies on the CT-value trajectories of Omicron BA1/BA2 infections. An earlier study conducted in March–May 2020 found that higher SARS-CoV-2 antibodies correlated with higher overall CT-values during infection (i.e. lower viral loads), but in this study SARS-CoV-2 antibodies and infection involved closely related/identical variants.^20^ Other studies observed no difference in CT-value trajectories after vaccination, regardless of vaccination status for Alpha, Delta, and Omicron^21,22^. Some studies have noticed an influence of cross-variant SARS-CoV-2 antibodies on patterns of viral reproduction. Puhach et al. (2022) ^23^ noticed that SARS-CoV-2 vaccination, which boosts spike ectodomain and spike S1, reduces viral load among individuals infected with the Delta variant during the first 5 days post-onset of symptoms^23^. Another study observed that individuals with at least five exposures, vaccine or infection, have lower viral load peak, meaning less shedding, compared to individuals with three or four exposures^24^. Of note, we observed distinct patterns in CT-values for milder and asymptomatic infections versus symptomatic infections confirming earlier findings from ^10,20,25^, who reported an association between lower CT values - or, conversely, higher viral loads - and more severe COVID-19 cases.

The interpretation of our study’s results is subject to several limitations. First, we analyzed the effect of pre-existing antibodies on Omicron BA.1/BA.2 outcomes, but we cannot assume that our results can be directly applied to other, more recent variants. Second, the use of dried blood spots in a final dilution of 1:40 for immunoglobin G antibodies reactive with the target antigens posed technical challenges in accurately quantifying antibody titers, particularly at extreme values, with antibodies above 67,000 remaining unassessed due to right-censoring. This may have limited our sensitivity to detect some correlations. Third, the analyses on infection risk included coprimary cases, and infection risk might therefore be overestimated. However, our results remained robust in sensitivity analysis excluding co-primary cases. Lastly, the duration of RT-PCR positivity and CT-value < 30 may have been slightly underestimated as the sampling frequency was reduced after the first week. These results should therefore be interpreted with some caution due to the sampling protocol, potentially leading to misinterpretation of viral CT-value kinetics. Therefore, the mean duration of RT-PCR positivity and CT-value <30 might be underestimated.

## Conclusion

Our study indicates that prior SARS-CoV-2 and beta seasonal hCoV antibodies provide protection against Omicron BA.1/BA.2 infection, but limited or no effect on symptom burden and CT-value trajectories. The results provide further insight on the relatively small, but not negligible role of these prior antibodies, especially when facing immune escape variants of SARS-CoV-2.

## Author Contributions

Conceptualization: P Bruijning-Verhagen. Coordination data collection: P Bruijning-Verhagen, I Westerhof. Lab analysis: R Sikkema, J van Beek. Data management: I Westerhof. Methodology: I Westerhof, P Bruijning-Verhagen. Investigation: I Westerhof. Formal analysis: I Westerhof. Visualization: I Westerhof. Writing—original draft: I Westerhof, P Bruijning-Verhagen. Writing— review and editing: I Westerhof, R Sikkema, G Rozhnova, J van Beek, M Koopmans, P Bruijning-Verhagen.

## Acknowledgements

We gratefully acknowledge all participants of the study.

## Supplementary Figures

**Supplementary Fig 1.** Correlation of cumulative high antibody levels for different coronaviruses measured at baseline. P-values: *** <0.001, ** <0.01, *<0.05, .<0.1.

**Supplementary Fig. 2** SARS-CoV-2 infection status (Red: Infected; Blue: Not infected) by baseline antibody levels for different coronaviruses. Boxplots represent the median with IQR. P-values: *** <0.001, ** <0.01, *<0.05.

**Supplementary Fig. 3.** Disease severity (Red: Symptomatic; Blue: Pauci-symptomatic; Green: Asymptomatic) by baseline antibody levels for different coronaviruses, among SARS-CoV-2 infected household members. Boxplots represent the median with IQR. P-values: **** <0.0001, *** <0.001, ** <0.01, *<0.05, .<0.1.

