## Supplementary figures and images for "The effect of pre-existing coronavirus antibodies on SARS-CoV-2 infection outcomes in exposed household members"

### Suppl. figure 1

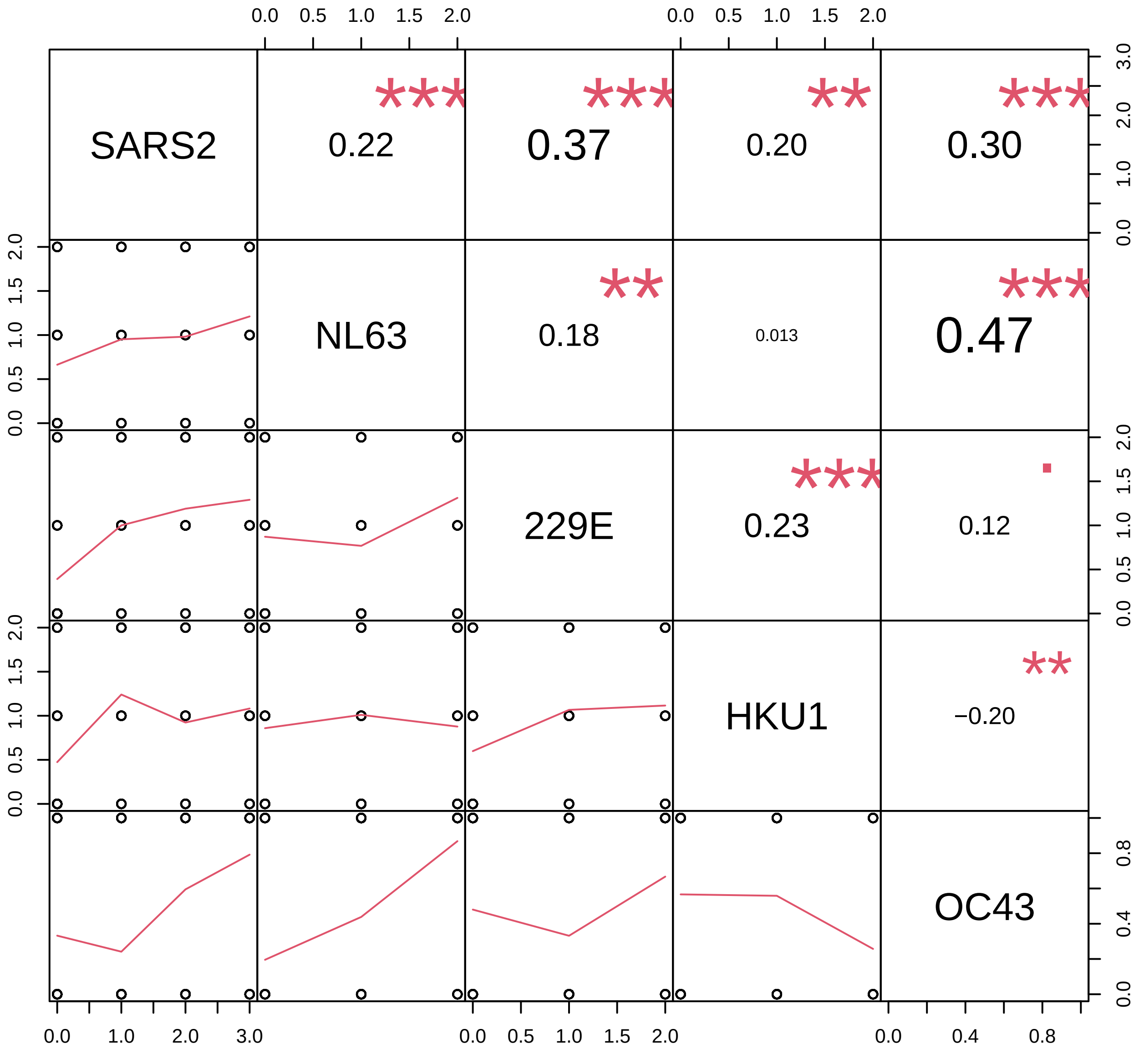

### Suppl. figure 2

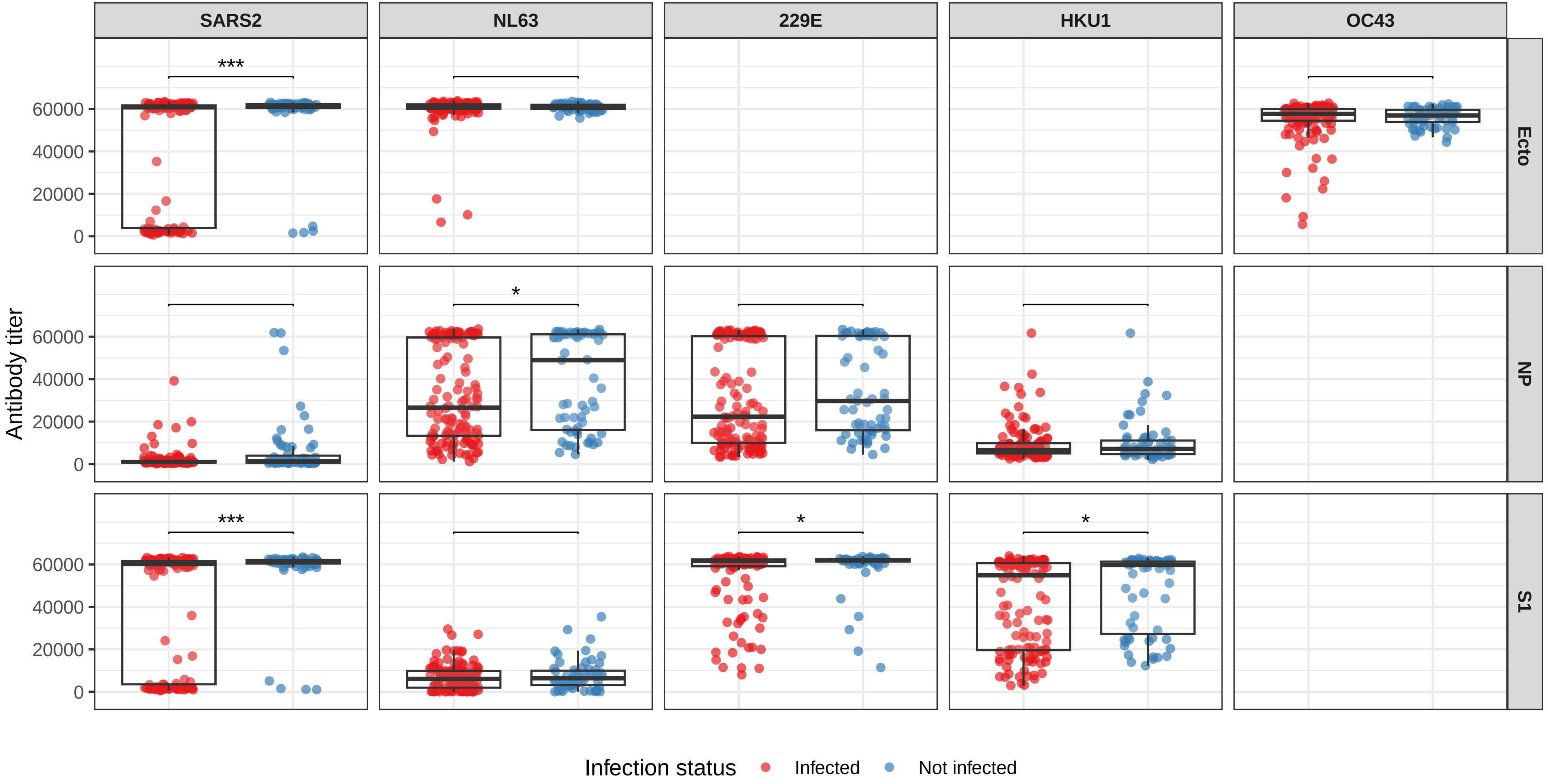

### Suppl. figure 3

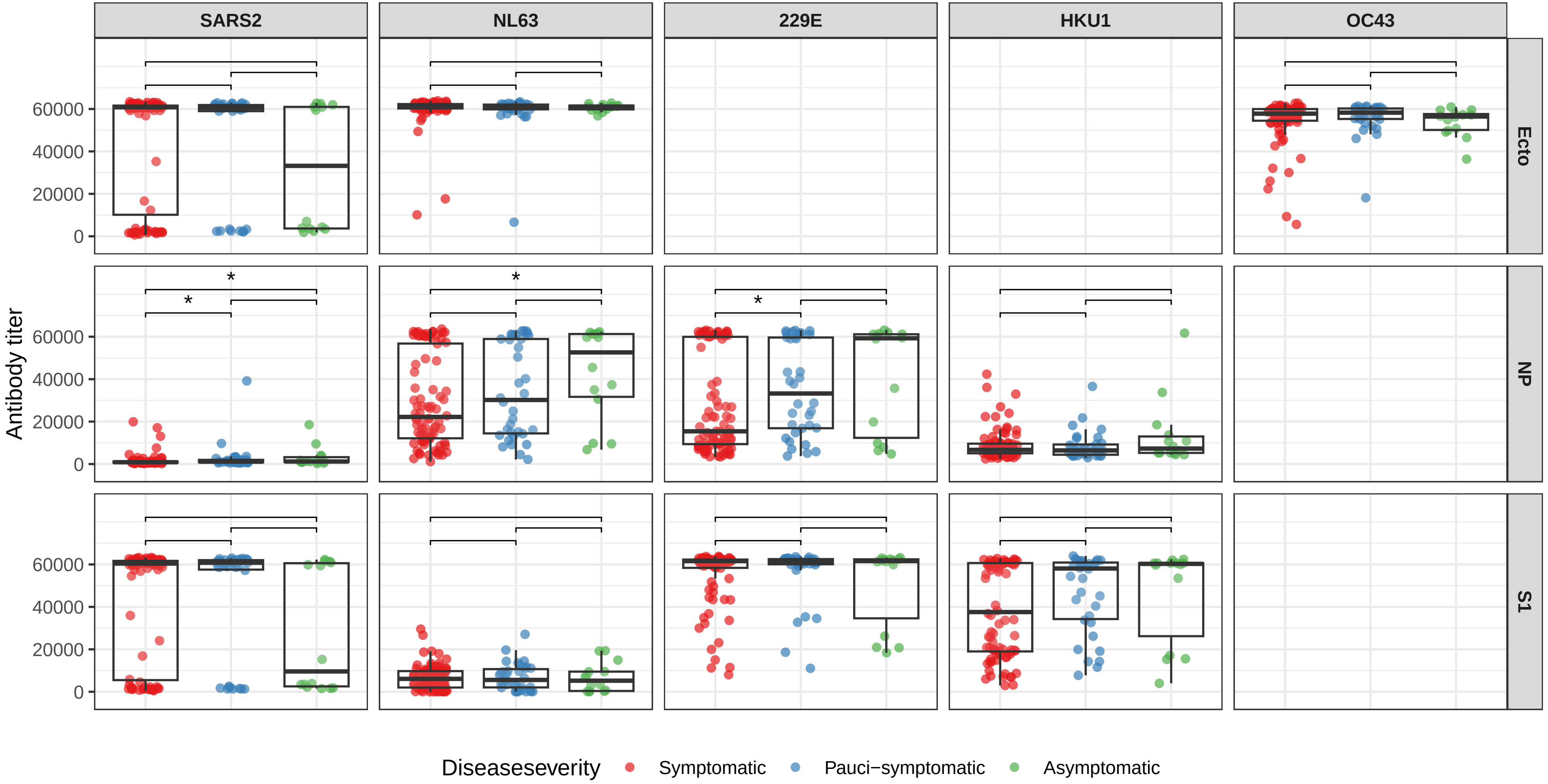
